# Prospective population-based observational study to estimate the incidence of T2DM in a metropolitan population in the north of Madrid (Spain) and to determine the effect of baseline glycaemic status through an explanatory Cox model. SPRINDIAP-1 study (Secondary PRevention of INcident DIAbetes in patients with Prediabetes)

**DOI:** 10.1101/2023.06.16.23291494

**Authors:** V Iriarte-Campo, C de Burgos-Lunar, J Mostaza, C Lahoz, J Cárdenas-Valladolid, P Gómez-Campelo, B Taulero-Escalera, FJ San-Andrés-Rebollo, F Rodriguez-Artalejo, MA Salinero-Fort, SPREDIA-2 Group

## Abstract

**Background:** T2DM (Type 2 Diabetes mellitus) is preceded by basal glycaemic states (BGS) such as normoglycaemia (NG) or pre-T2DM, including impaired fasting glucose (IFG); impaired glucose tolerance (IGT) or both (IFG-IGT). A better understanding of the role of pre-T2DM in the progression to T2DM may help in the prevention of T2DM in patients with pre-T2DM.

**Methods:** Population-based cohort study to estimate the incidence of T2DM according to BGS and to study the adjusted effect of BGS on progression to T2DM using a Cox model (main model (MM) with reference category NG and sensitivity analysis (SA) in patients with pre-T2DM and reference IFG).

**Results:** 1,209 patients aged 45-74 years (median follow-up=7.26 years). The crude T2DM incidence rate for the whole population was 11.21 per 1000 person-years (95%CI=9.09-13.68), 5.60 (95%CI=3.55-8.41) in patients with NG and 16.28 (95%CI=12.78-20.43) in patients with preT2DM. In both models, the significant variables showing risk of progression to T2DM were IGT BGS (MM: Hazard ratio HR=4.30; 95%CI=1.96-9.44; AS: HR=2.66, 95%CI=1.28-5.56) and IFG-IGT (MM: HR=3.71, 95%CI=1.97-6.99; AS: HR=2.45, 95%CI=1.41-4.23), and obesity (MM: HR=2.36, 95%CI=1.15-4.83; AS: HR=2.97, 95%CI=1.18-7.45). Being on diuretic treatment was a protective factor (MM: HR=0.47 CI95%=0.23-0.96; SA: HR=0.41, CI95%=0.19-0.92), as was, in SA only, self-perceived health status in the categories of: Very good (HR: 0.19, CI95%=0.06-0.67); Good (HR: 0.35, CI95%=0.13-0.96) and Fair (HR: 0.31, CI95%=0.11-0.93).

**Conclusions:** Our T2DM incidence rates are in line with other Spanish studies. In people with NG or preT2DM, EBG (IGT and IFG-IGT) and obesity increase the risk of progression to T2DM and being on diuretics is a protective factor as is fair to very good self-perceived health in patients with preT2DM.

## Background

Diabetes mellitus (DM) is a rising global health problem that affects approximately 8.3% of the adult population worldwide, 90% of whom have type 2 diabetes mellitus (T2DM) (1). More concretely, between 2019 and 2045, the number of adults with T2DM is expected to increase from 463.0 million to 700.2 million, implying an increase in annual costs related to its management during this period from USD 760.3 billion to USD 845.0 billion (2).

From the patients’ perspective, patients with T2DM have, on average, 2.2 times higher societal costs, such as productivity losses and informal care costs, and experience lower quality of life than people without the disease (3). Even if patients suffer from 2 or more diabetes-related complications, the total social costs are 4.8 times higher than those without them (p<0.05) (3). T2DM complications include cardiovascular disease, retinopathy, neuropathic pain, nephropathy nontraumatic lower limb amputation and cancer, among others (4,5), and some may appear as early as the time of diagnosis of the disease (6).

Disease frequency studies such as prevalence and incidence studies describe the impact of a health problem on a given population (7,8). In addition, when we estimate the incidence of T2DM, we have more reliable information on the predictive factors involved in disease progression (9), which is also useful for assessing the success of public health campaigns for the prevention of T2DM (4). Therefore, the International Diabetes Federation (IDF) recommends carrying out incidence studies in different populations (5).

T2DM is often preceded by a long period of prediabetes (pre-T2DM) —a state of glucose dysregulation between normoglycemia (NG) and T2DM— that increases not only the risk of progression to T2DM (5,9–12) but also the risk of having T2DM complications (20) or suffering cardiovascular events such as myocardial infarction, stroke, or death from cardiovascular causes(12). The pre-T2DM phenotype can be characterised by blood glucose levels, including FPG and 2 h-OGTT glucose (5,13,14). Several studies have been carried out in our country in which pre-T2DM or altered levels of FPG have been identified as predictors of progression to T2DM(4,5,9,15). On the other hand, some studies have demonstrated that early identification of pre-T2DM when accompanied by lifestyle interventions may reduce the risk of developing T2DM (16) with a prolonged effect in reducing the risk of complications and comorbidities as well as improving intermediate health outcomes(17–21) such as improved health-related quality of life, decreased health-system cost and potentially decreased cardiovascular risk (22).

Spanish studies on T2DM incidence have been carried out either on a national scale (5,8,23) or in different parts of Spain (4,9,13) with mixed results, and it is possible that the north‒south axis is reproduced in T2DM as in other diseases (obesity, hypertension, hypercholesterolemia, coronary artery disease)(24,25), although this needs to be confirmed with current data. In addition, no studies of T2DM incidence rates per 1,000 person-years have been carried out in Madrid(26).

Regarding predictors of progression to T2DM, only 4 studies (4,5,9,13) established baseline glycaemic status (BGS) by classifying patients at baseline as persons with normoglycaemia (NG) or patients with prediabetes, differentiating within the latter between patients with isolated impaired fasting glucose (IFG), patients with isolated impaired glucose tolerance (IGT) or patients with both (IFG-IGT). However, none has explained the specific role of BGS in the progression to T2DM with patients’ self-perception of health as an independent variable along with cardiometabolic variables.

The aims of the SPRINDIAP-1 study are to estimate the incidence of T2DM and to determine the effect of BGS, after controlling for the effect of all other potential confounding factors, on progress to T2DM. In doing so, we aim to contribute to a better understanding of the specific role of pre-T2DM in the progression to T2DM and to better management of patients with prediabetes in the context of secondary prevention strategies for T2DM in Madrid.

## Methods

### Study design

The SPRINDIAP-1 study is a population-based prospective cohort study conducted in Madrid (Spain) of persons with NG and patients with prediabetes (isolated IFG, isolated IGT or IFG-IGT) to estimate the incidence of T2DM after 7.26 years of follow-up and to determine the specific adjusted effect of BGS on progression to T2DM.

### Data source and study population

SPRINDIAP-1 is the follow-up study of patients free of DM at baseline included in a larger project funded by the Fondo de Investigación Sanitaria of the Instituto de Salud Carlos III in 2015 (registration number PI1500259), whose baseline screening (SPREDIA-2) was published in 2015 (26).

The T2DM criteria (27) used to exclude patients at baseline were as follows: FPG > 126 mg/dL and 2 h OGTT glucose > 200 mg/dL; FPG > 126 mg/dL and 2 h OGTT glucose with data not available; 2 h OGTT glucose > 200 mg/dL; HbA1C > 6.5% or patients taking any antidiabetic medication. See the patient flow chart in Figure 1 for more information.

**Figure 1.**
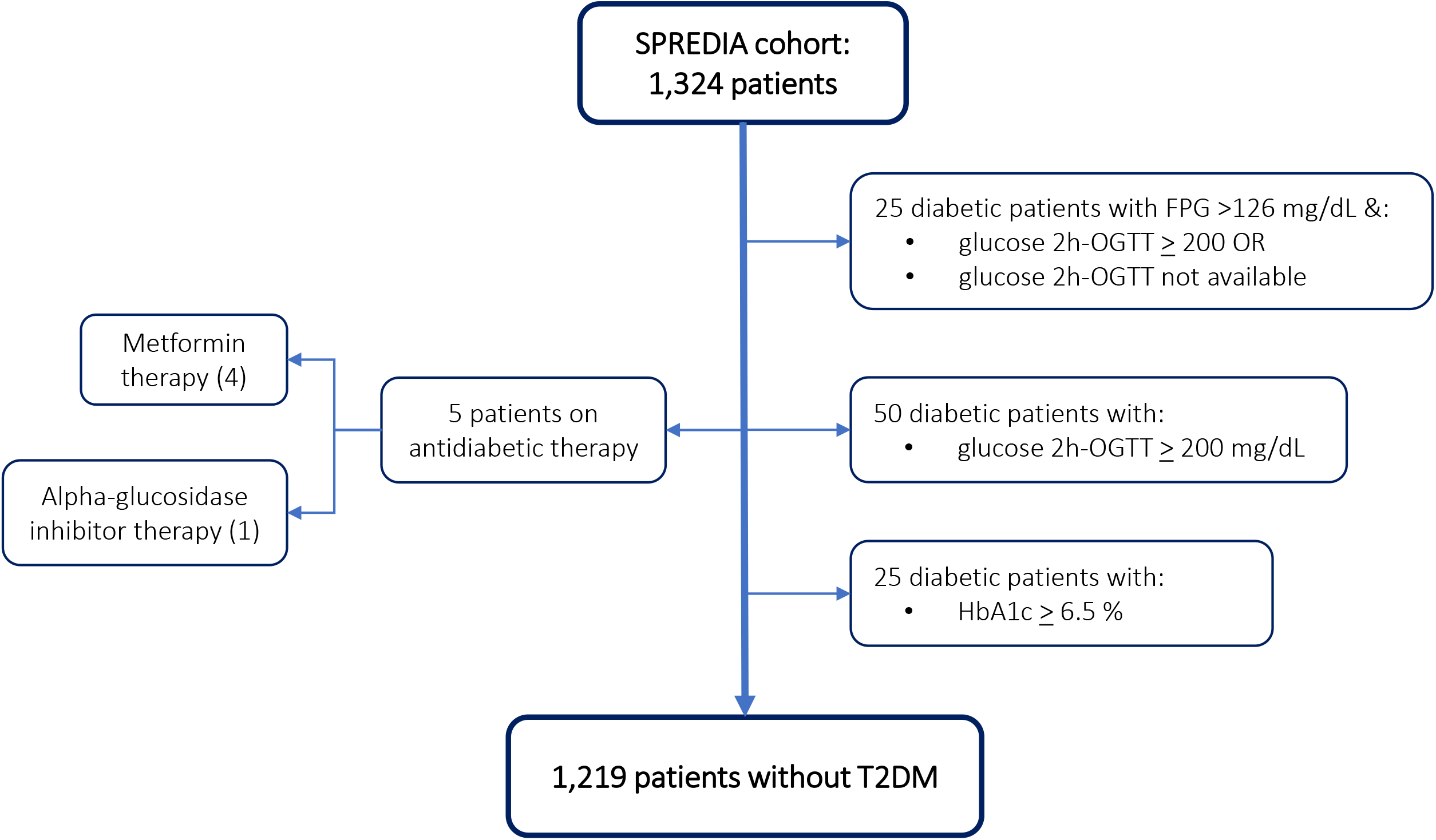
SPRINDIAP-1 study flow chart of patients and their segmentation by baseline glycaemic status.

Clinical data to determine SPRINDIAP-1 outcomes in this population (development of T2DM) were obtained from the Madrid Primary Care Electronic Health Record (PC-EHR). Mortality data were obtained from a death certificate database available from the Spanish National Institute of Statistics.

### Classification of the study population with pre-DM

According to the American Diabetes Association (ADA) criteria (27), SPRINDIAP-1 patients with prediabetes were characterised into 3 mutually exclusive subgroups defined as follows:

- Patients with isolated IFG: FPG levels from 100 mg/dL to 125 mg/dL (both included) and 2 h-OGTT glucose < 140 mg/dL.
- Patients with isolated IGT: FPG < 100 mg/dL and 2 h-OGTT glucose between 140 mg/dL and 199 mg/dL (both included).
- IFG-IGT patients: FPG levels from 100 mg/dL to 125 mg/dL (both included) and 2 h-OGTT glucose between 140 mg/dL and 199 mg/dL (both included).

In addition, patients with FPG < 100 mg/dL and a 2 h-OGTT glucose < 140 mg/dL were considered normoglycaemic. The ADA criteria were chosen because they have been shown to have a lower false positive rate and higher sensitivity (8) in determining the BGS of patients.

Thirty-six patients could not be categorised by their BGS, either because 2 h-OGTT glucose data were not available or because their FPG and 2 h-OGTT glucose values did not correspond to any category established by the ADA criteria. Therefore, their characterisation was determined at the discretion of the study Steering Committee (see supplementary material).

### T2DM

T2DM was defined as FPG ≥126 mg/dL or HbA1c ≥6.5% (27) or use of hypoglycemic medication during the follow-up period according to PC-EHR records during the follow-up period. As only annual data were available for both blood tests and first antidiabetic treatment, the date of diagnosis of T2DM was arbitrarily set to June 1 of the year in which either approach suggested a case of T2DM. Thus, to avoid negative follow-up periods in subjects whose year of entry into the study coincided with the year in which a case of T2DM was identified, patients should have been recruited before 31 May. For further information on the algorithms used to identify incident cases of T2DM, see the Supplemental material.

Regarding follow-up time, for living patients who did not develop T2DM, the time at risk was the difference between the date of entry into the SPREDIA cohort and the SPRINDIAP-1 study end date (31 December 2019). In case of death or onset of T2DM, follow-up time was calculated until the earliest of these two dates.

### Statistical analysis

First, we analysed the distribution of baseline characteristics of the study population by BGS category. For each variable, we also analysed whether the differences in this distribution were statistically significant compared with the reference category. To do this, we used the nonparametric Kruskal‒Wallis test for continuous variables and the chi-squared test for linear trend for categorical variables, as they were ordinal variables. Both the distribution and comparisons are shown in Table 1.

**Table 1.**
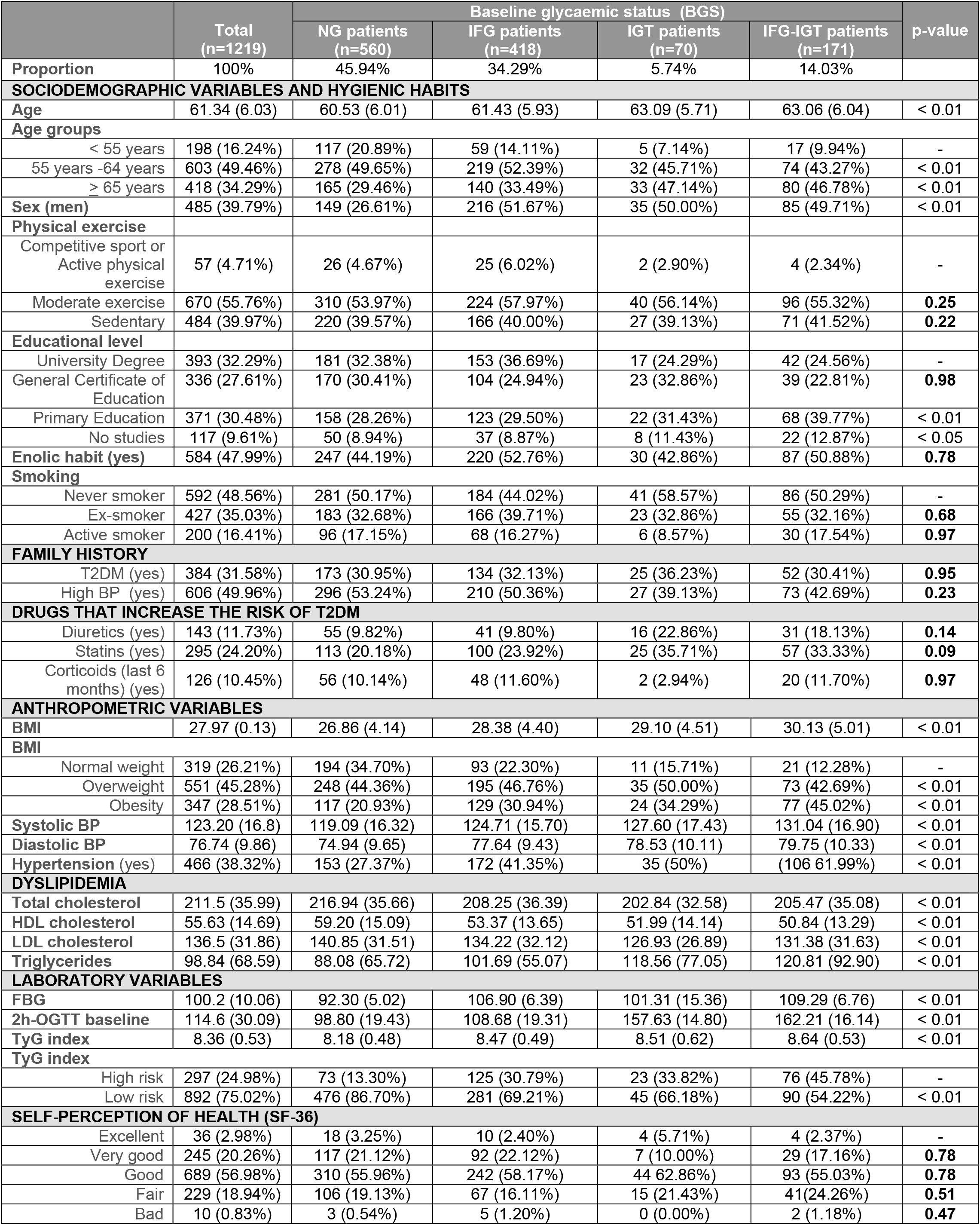
Baseline characteristics of the SPRINDIAP-1 population. Data are means ± SD or count (%). BMI: Body mass Index in kg/m^2^; BP Blood pressure in mmHg; CHD: coronary heart disease; CVD: cerebrovascular disease; CGV: Coefficient of glycaemic variation; FBG: Fasting blood glucose; HDL cholesterol in mg/dL; IFG: patients with isolated impaired fasting glucose; IGT patients with isolated impaired glucose tolerance; IFG-IGT: patients with both; LDL cholesterol in mg/dL; NG: patients with normoglycemia; OGTT: Oral Glucose Overload Test; SF-36: SF-36 Self-perceived health status questionnaire; Total cholesterol in mg/dL; Triglycerides in mg/dL; TyG: Triglyceride glucose index

#### T2DM incidence

The cumulative incidence of T2DM was calculated, together with its 95% confidence interval (CI95%), as the total number of new cases of T2DM in the analysed population stratum divided by the total number of persons at risk in that stratum. The raw incidence rate (expressed as the number of cases per 1,000 person-years) was calculated as above, but the total follow-up time of all persons at risk of developing T2DM was included in the denominator, and the result was multiplied by 1,000. The incidence rate ratio was calculated by dividing the incidence rate of patients with prediabetes, isolated IFG, isolated IGT or IFG-IGT by the incidence rate of patients with NG.

On the other hand, the standardised incidence rate by BGS category was calculated both in the total population and by age, differentiating between males and females using the direct method with an internal standard population (for further details on the calculation methods for this purpose, see the supplementary material).

#### Survival analysis

As a first approximation to the survival model, we plotted the Kaplan‒Meier curve for the subtypes of BGS and tested the hypothesis that the groups compared had equal survival. We verified that the Kaplan‒Meier curves of the isolated IGT and IFG-IGT patients crossed (see Figure 3), so we used the Breslow test to compare them (p value <0.01).

Cox proportional hazards explanatory model was carried out, with BGS as the main independent variable and progression to T2DM as the dependent variable. Previously, we estimated the cumulative individual survival probability over time or the fraction of event-free patients with Kaplan‒Meier curves for the main variable.

To estimate the instantaneous risk of progression to T2DM from isolated IFG, isolated IGT, IFG-IGT, and NG, we calculated the hazard ratio (HR) and 95% CI of each category of BGS, with NG as the reference, controlling for the potential confounding effect of the other factors included in the multivariable Cox model (see Table 3). We also performed a sensitivity analysis (the same models applied only to the population with prediabetes, with isolated IFG as the reference category). The likelihood ratio test was used to check the goodness of fit, and a Schoenfeld residual test was used to check the proportional risk assumption of the Cox model (see the Supplementary Appendix).

We included the following SPREDIA-2 baseline variables (26) as independent variables in both the Cox model and the sensitivity analysis: sociodemographic (age, sex), cardiometabolic risk (alcohol consumption; smoking; family history of T2DM and hypertension; level of physical activity; body mass index; diagnosis of hypertension; plasma levels of total, HDL and LDL cholesterol; plasma levels of triglycerides and triglyceride-glucose ratio). The triglyceride-glucose ratio is a simple surrogate estimate of insulin resistance (28) and was calculated using the formula ln[fasting triglycerides (mg/dL) × fasting glucose (mg/dL)/2] (28). Hypertension was defined as systolic blood pressure ≥140 mmHg and/or diastolic blood pressure ≥90 mmHg or treatment with antihypertensive drugs, and body mass index (BMI) was categorised as normal weight (< 24.9 kg/m2), overweight (25.0 - 29.9 kg/m2) or obese (> 30.0 kg/m2). Treatments that increase the risk of T2DM (diuretics, statins or corticosteroids) and self-perceived health using the SF-36(28) were also taken into account.

### Ethics

The PI1500259 project was approved in 2015 by the Research Ethics Committee of the Hospital Universitario Ramón y Cajal (Madrid), approval that included future studies in which its patients would be followed up. Specifically, the SPRINDIAP-1 study follows up patients who had prediabetes at baseline in the PI1500259 project. Since SPRINDIAP-1 clinical data have been obtained from the patients’ PC-EHR (secondary data), our study was exempt from obtaining the signature of a new informed consent form from patients.

### Data processing, analysis, and access

Processing the study data, descriptive analysis, cumulative and rate incidence, and multivariate analysis by Cox model were carried out with R version 2022.12.0 Build 353. Age-standardised rates of T2DM were determined with Microsoft Excel 365. Datasets generated and/or analysed during the current study are available from the corresponding author on reasonable request.

## Results

### Baseline population

The SPRINDIAP-1 study population comprised 1,209 patients aged 45-74 years with a median follow-up of 7.26 years. The flow chart of individuals from the original SPREDIA-2 cohort to those free of T2DM is shown in Figure 1.

Table 1 shows the descriptive analysis stratified by BGS categories of all variables in the SPRINDIAP-1 study. Older age groups and those with less education were significantly more likely to have IGT, isolated or combined with IFG. The same was true for most anthropometric variables, dyslipidaemia and perceived fair or bad health according to the SF-36 questionnaire. More detailed figures on the distribution of BGS by age and sex can be found in the supplementary material.

### T2DM incidence

The raw incidence rate of T2DM was 11.21 cases per 1,000 person-years (95% CI, 9.09 -13.68) for the whole population, 5.60 cases per 1,000 person-years (95% CI, 3.55 - 8.41) for patients with NG and 16.28 cases per 1,000 person-years (95% CI, 12.78 - 20.43) for all patients with prediabetes. The rate ratio between patients with prediabetes and NG was 2.91 (95% CI, 1.80 -4.86). When stratified by type of prediabetes, as described in the supplementary material, the raw incidence rates of T2DM were as follows: 10.71 cases per 1,000 person-years for patients with isolated IFG (95% CI, 7.32 - 15.11); rate ratio between patients with isolated IFG and NG was 1.91 (95% CI, 1.08 – 3.42); 26.91 cases per 1,000 person-years for patients with isolated IGT (95% CI, 13.90 - 47.00); rate ratio between patients with isolated IGT and NG: 4.80 (95% CI, 2.18 – 10.06) and 27.00 cases per 1,000 person-years for patients with IFG-IGT (95% CI, 18.22 -38.55; rate ratio between patients with IFG-IGT and NG: 4.82 (95% CI, 2.71 – 8.69). There were no statistically significant differences in the raw incidence rate ratio of T2DM by either sex or age (see supplementary material for more information).

The standardised T2DM incidence rates showed similar results to the crude rates as can be seen in the supplementary material.

The Sankey plot in Figure 2 plots the flow of patients between the four BGS categories and incident T2DM after 7.26 years of follow-up, as well as the cumulative incidence of T2DM by stratum.

**Figure 2.**
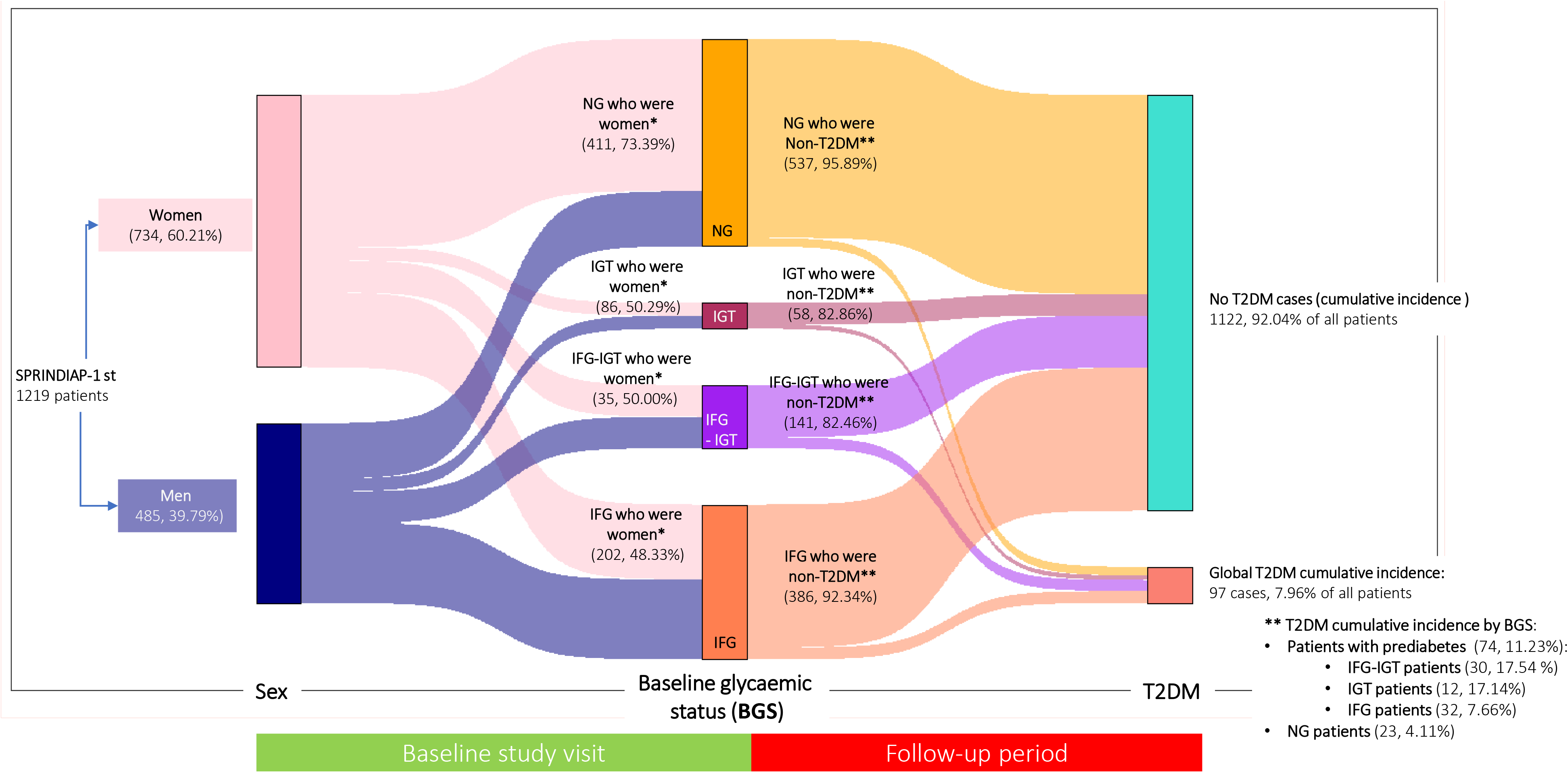
Sankey plot representing the progression of study patients to T2DM cases, once categorised by sex and then by BGS (Figures in count and %) IFG: patients with isolated impaired fasting glucose; IGT patients with isolated impaired glucose tolerance; IFG-IGT: patients with IFG and IGT; NG: patients with normoglycaemia * Complementary values in men: 26.61% with NG; 49.71% with IGT; 50% with IFG-IGT and 51.67% with IFG.; ** Complementary T2DM values are detailed in T2DM cumulative incidence by BGS

The crude incidence rate ratio of T2DM in subjects with prediabetes to patients with NG was close to three (2.91; 95% CI, 1.80-4.86; p value = 0.001), with small differences between men (3.05; 95% CI, 1.28-8.81; p value = 0.005) and women (2.76; 95% CI, 1.51-5.26). When analysing age subgroups, rate ratios below 2 did not reach statistical significance, as shown in Table 2. The age subgroup with the highest rate was men aged 55-64 years, with a rate ratio close to 10 (9.62; 95% CI, 1.55-397.9: p value = 0.003), which was three times higher than that for women in the same age group (3.27; 95% CI, 1.38-8.59; p=0.003).

**Table 2.** Raw T2DM rates by baseline glycaemic status and age group in males and females (Pre-T2DM vs NG)

|  |  | Pre-T2DM at baseline (n=659) |  |  |  | NG: Normoglycemia at baseline (n=560) |  |  |  |  |  |
| --- | --- | --- | --- | --- | --- | --- | --- | --- | --- | --- | --- |
|  |  | n | T2DM cases | p-y | T2DM incidence per 1000 p-y (95% CI) | n | T2DM cases | p-y | T2DM incidence per 1000 p-y (95% CI) | Raw rate ratio (Pre-T2DM /NG) (95% CI) | p-value |
| **All** | **<55 years old** | 81 | 6 | 567.62 | 10.57 (3.88-23.01) | 117 | 5 | 878.40 | 5.69 (1.85-13.28) | 1.86 (0.47-7.69) | 0.321 |
|  | **55-64 years old** | 325 | 41 | 2250.50 | 18.22 (13.07-24.7) | 278 | 9 | 2025.70 | 4.44 (2.03-8.43) | 4.10 (1.96-9.60) | <0.001 |
|  | **≥65 years old** | 253 | 27 | 1728.30 | 15.62 (10.30-22.73) | 165 | 9 | 1201.00 | 7.49 (3.43-14.22) | 2.08 (0.95-5.04) | 0.049 |
|  | **Total** | **659** | **74** | **4546.42** | **16.3 (12.78-20.43)** | **560** | **23** | **4105.1** | **5.6 (3.55-8.41)** | **2.91 (1.80-4.86)** | **<0.001** |
| **Men** | **<55 years old** | 53 | 5 | 360.88 | 13.86 (4.50-32.33) | 40 | 1 | 306.60 | 3.26 (0.08-18.17) | 4.25 (0.48-200.9) | 0.177 |
|  | **55-64 years old** | 158 | 21 | 1081.70 | 19.41 (12.02-29.68) | 69 | 1 | 495.70 | 2.02 (0.05-11.24) | 9.62 (1.55-397.9) | 0.003 |
|  | **≥65 years old** | 125 | 13 | 842.40 | 15.43 (8.22-26.39) | 40 | 4 | 269.40 | 14.85 (4.05-38.02) | 1.04 (0.32-4.38) | 0.984 |
|  | **Total** | **336** | **39** | **2284.98** | **17.1 (12.14-23.33)** | **149** | **6** | **1071.7** | **5.6 (2.06-12.19)** | **3.05 (1.28-8.81)** | **0.005** |
| **Women** | **<55 years old** | 28 | 1 | 206.74 | 4.84 (0.12-26.95) | 77 | 4 | 571.80 | 7.00 (1.91-17.91) | 0.69 (0.01-6.99) | 0.814 |
|  | **55-64 years old** | 167 | 20 | 1168.80 | 17.11 (10.45-26.43) | 209 | 8 | 1530.00 | 5.23 (2.26-10.30) | 3.27 (1.38-8.59) | 0.003 |
|  | **≥65 years old** | 128 | 14 | 885.90 | 15.80 (8.64-26.52) | 125 | 5 | 931.60 | 5.37 (1.74-12.53) | 2.94 (1.01-10.44) | 0.032 |
|  | **Total** | **323** | **35** | **2261.44** | **15.48 (10.78-21.53)** | **411** | **17** | **3033.4** | **5.6 (3.27-8.97)** | **2.76 (1.51-5.26)** | **<0.001** |

Pre-T2DM: Prediabetes; p-y: person-years; T2DM: Type 2 Diabetes Mellitus

### Survival model

The Kaplan‒Meier curves showed that NG and isolated IFG were significantly more likely to remain free of T2DM than isolated IGT or IFG-IGT (p value <0.01) (Figure 3). Table 3 describes the adjusted HR of the four BGS categories, from a model adjusted only for age and sex (model 1) to a fully adjusted model (model 5), distinguishing between the main Cox model (run in the whole population, with NG as the reference category) and the sensitivity analysis (model in patients with prediabetes, with isolated IFG as the reference category). The final model selected is number five.

**Figure 3.**
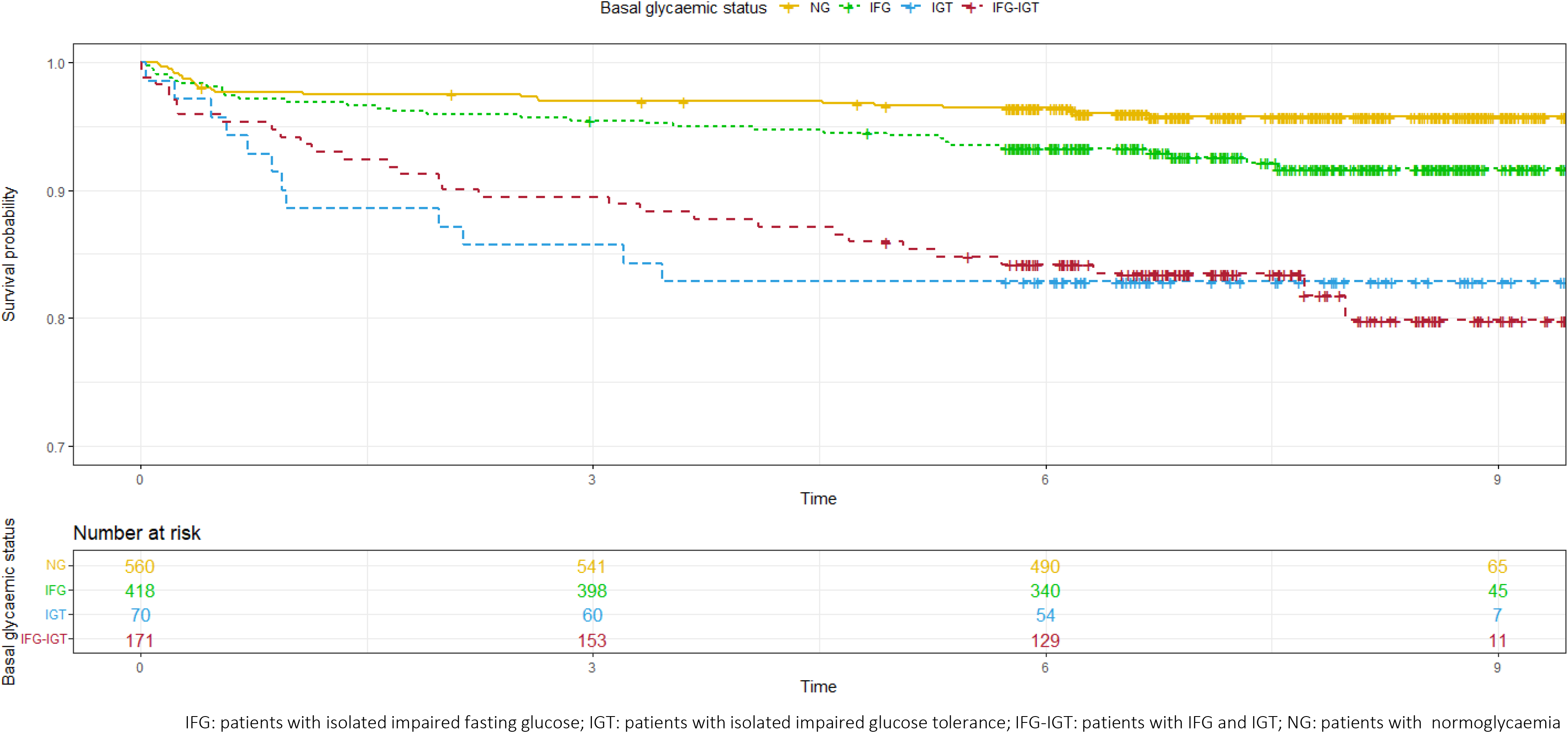
Probability of remaining free of T2DM Kaplan-Meier survival analysis. Differences by baseline glycaemic status.

**Table 3-A.** Cox proportional hazards model with all the study patients.

| Variable | Patients | MODEL 1 |  |  | MODEL 2 |  |  | MODEL 3 |  |  | MODEL 4 |  |  | MODEL 5 (final) |  |  |
| --- | --- | --- | --- | --- | --- | --- | --- | --- | --- | --- | --- | --- | --- | --- | --- | --- |
|  |  | HR | lower .95 | upper .95 | HR | lower .95 | upper .95 | HR | lower .95 | upper .95 | HR | lower .95 | upper .95 | HR | lower .95 | upper .95 |
| Baseline glycaemic status | NG | 1 | 1 | 1 | 1 | 1 | 1 | 1 | 1 | 1 | 1 | 1 | 1 | 1 | 1 | 1 |
|  | IFG | **1.85** | **1.07** | **3.20** | **1.84** | **1.06** | **3.23** | 1.74 | 0.99 | 3.08 | 1.61 | 0.89 | 2.91 | 1.61 | 0.89 | 2.90 |
|  | IGT | **4.51** | **2.22** | **9.17** | **4.27** | **2.02** | **8.91** | **4.05** | **1.91** | **8.58** | **4.29** | **1.95** | **9.42** | **4.30** | **1.96** | **9.44** |
|  | IFG-IGT | **4.53** | **2.59** | **7.94** | **4.63** | **2.62** | **8.18** | **3.89** | **2.16** | **7.02** | **3.69** | **1.96** | **6.96** | **3.71** | **1.97** | **6.99** |
| Likelihood ratio test |  | 36.88 on 6 df, p< 0.01 |  |  | 57.22 on 14 df, p< 0.01 |  |  | 68.21 on 21 df, p< 0.01 |  |  | 79.97 on 30 df, p< 0.01 |  |  | 79.93 on 27 df, p< 0.01 |  |  |

**Table 3-B.** Cox proportional hazards model in patients with pre-T2DM (sensitivity analysis)

| Variable | Patients | MODEL 1 |  |  | MODEL 2 |  |  | MODEL 3 |  |  | MODEL 4 |  |  | MODEL 5 (final) |  |  |
| --- | --- | --- | --- | --- | --- | --- | --- | --- | --- | --- | --- | --- | --- | --- | --- | --- |
|  |  | HR | lower .95 | upper .95 | HR | lower .95 | upper .95 | HR | lower .95 | upper .95 | HR | lower .95 | upper .95 | HR | lower .95 | upper .95 |
| Baseline glycaemic status | IFG | 1 | 1 | 1 | 1 | 1 | 1 | 1 | 1 | 1 | 1 | 1 | 1 | 1 | 1 | 1 |
|  | IGT | **2.47** | **1.26** | **4.81** | **2.15** | **1.06** | **4.35** | **2.23** | **1.10** | **4.53** | **2.69** | **1.29** | **5.63** | **2.66** | **1.28** | **5.56** |
|  | IFG-IGT | **2.49** | **1.51** | **4.13** | **2.58** | **1.54** | **4.32** | **2.29** | **1.35** | **3.86** | **2.39** | **1.38** | **4.14** | **2.45** | **1.41** | **4.23** |
| Likelihood ratio test |  | 16.00 on 5 df, p< 0.01 |  |  | 35.93 on 13 df, p< 0.01 |  |  | 44.70 on 20 df, p< 0.01 |  |  | 56.48 on 29 df, p< 0.01 |  |  | 55.08 on 26 df, p< 0.01 |  |  |

**Model 1:** Baseline glycaemic status, age (as a continuous variable), gender (ref cat: women) and interaction of age and gender. **Model 2:** Model 1 plus Enolic habit (ref cat: no), Smoking (reference category: Never Smoker vs Ex-smoker and Active smoker), Self-perception of health by SF-36 questionnaire (ref cat: Excellent vs Very good, Good, Fair and Bad), Family history of T2DM (ref cat: no) and Family history of Hypertension (ref cat: no). **Model 3:** Model 2 plus Physical exercise (ref cat: Competitive sport or Active physical exercise vs Moderate exercise and Sedentary), Educational level (ref cat: No studies vs Primary Education, General Certificate of Education and University Degree) and Body Mass Index (ref cat: Normal weight vs Overweight and Obesity). **Model 4:** Model 3 plus Hypertension (ref cat: no), Complete lipid profile (Total cholesterol, HDL-cholesterol, LDL-cholesterol and Triglyceride levels in blood, all as quantitative variables), Exposure to diuretics (ref cat: no), Exposure to statins (ref cat: no), Exposure to corticoids within last 6 months (ref cat: no) and Glycaemia-Triglyceride Index as quantitative variable. **Model 5: Final model.** Comprises all variables in model 4 with the exception of the variable Educational level, extracted from the model to obtain a non-significant Schoenfeld residual test for the overall model, thus fulfilling the Cox model assumption that HR is constant over time.

In both models, the variables that were significant as risk factors for progression to T2DM were isolated IGT and IFG-IGT categories (Table 3), obesity (HR=2.36, 95% CI: 1.15 - 4.83 in the main model and HR= 2.97, 95% CI: 1.18 - 7.45 in sensitivity analysis) and being on diuretics (HR=0.47, 95% CI: 0.23 - 0.96 and HR=0.41, 95% CI: 0.19 - 0.92, respectively). Self-perceived health status was shown to be a protective factor only in the sensitivity analysis: very good (HR: 0.19, 95% CI: 0.06 - 0.67); good (HR: 0.35, 95% CI: 0.13 - 0.96) and even fair (HR: 0.31, 95% CI: 0.11 - 0.93), compared with excellent as the reference category.

Finally, we verified compliance with the proportional hazards assumption of the Cox model in the main model and in the sensitivity analysis with the Schoenfeld residuals test. Both the test of main and sensitivity model (p > 0.05) as well as of the main variable in each model (p > 0.05) were significant, thereby complying with the assumption.

## Discussion

This study reports the incidence rate of T2DM and the specific effect of BGS on progression to T2DM after 7.26 years of follow-up in a metropolitan population from the north of the city of Madrid.

Regarding the first objective, it is striking that the crude incidence rate obtained in our study (11.21 cases/1,000 person-years) is almost in the middle of the Spanish north‒south axis, represented by the incidence rates described by Urrutia et al.(9) in 2021 in Basque Country (6.56 cases/1,000 person-years, mean baseline age 52.3 years, 43.7% male, 87.55% of patients with NG and 23.30% obese) and that of the Pizarra study (13) carried out in Malaga (19.1 cases/1,000 person-years, mean age of 45.0 ± 13.4 years, 35.29% male, 69.30% of patients with NG and 32.80% obese), published one year later. Another relevant finding is the coincidence of our raw incidence rate with that of the Asturias study (4), although both studies present very different study populations in terms of the number of patients (630 vs 1,219 of our population), their age (51.5 ± 12.4 years vs 61.34 ± 6.03 years), the distribution of patients by BSG (> 80% of patients with NG vs 45.94%) or the percentage of obese patients (23.49% vs 28.51%). Although the patients in our study had a higher risk of developing diabetes at baseline than in the Asturias study (higher proportion of obese patients, older age, lower proportion of patients with normoglycaemia), the long-term incidence of diabetes was similar to that in the Asturias study conducted in 2006. One explanation for this apparent paradox could be the better control of diabetes risk factors in the last decade, as reported in several national(29,30) and international studies(31,32).

If we compare our results on the crude incidence rate of T2DM with those obtained in Spanish national studies, our rate is significantly higher. The Spanish national study (7) showed an incidence rate of 8.5 cases/1,000 person-years in a population with 87.80% of patients with NG, a mean age of less than 48 years and 26.20% of obese patients. Regardless of the difference in age and prevalence of obesity between our study and the Spanish national study (7), it should be noted that the definition of incident diabetes was not the same, which may influence the results.

In addition, two studies carried out in populations of workers, who are therefore healthier and younger than the general population, showed even lower incidence rates. Thus, Huerta et al. published in 2013 (25) a rate of 7.5 cases/1,000 person-years in men and 4.32 cases/1,000 person-years in women, with a mean age of approximately 50 years, and Vazquez et al. published in 2019 (17) figures of 5.0 cases per 1,000 person-years, with 88.25% of patients with NG and a mean age of 36.4 ± 10.4 years. These figures, which are lower than those found by us, are reasonable because of the bias in the sample.

Regarding the results obtained in the Cox model, note that 2 of the 3 BGS prediabetes categories (isolated IGT and IFG-IGT) were considered risk factors for progression to T2DM, as was highlighted previously (6,7,11,29). However, isolated IFG did not reach statistical significance in contrast with other studies(33–35).

Obesity was found to be a risk factor, in line with the results of many other studies (6,11,15,25,29), although no studies found that diuretic treatment was a protective factor for T2DM. In contrast, some studies have shown an increased risk of DM in patients using thiazide diuretics(36,37). However, data from the Atherosclerosis Risk in Communities (ARIC) study(38) do not show a significant increase in the risk of incident DM with diuretics (HR = 0.91, 95% CI, 0.73 - 1.13). The association between diuretics and incident DM is probably due to hypertension rather than the intrinsic long-acting effect of diuretics. The use of diuretics in subjects for reasons unrelated to hypertension or heart failure could explain weight loss as a reason for the protective effect on incident DM.

It is worth noting that 3 of the 4 categories of the SF-36 questionnaire (very good, good and fair) were found to be protective factors in the sensitivity analysis. It is plausible that patients with better health perceptions after receiving their BGS results may have adopted healthier diet and exercise behaviors in their lives, and this may be one of the explanations for being a protective factor against diabetes.

Hypertension did not show an independent association with the development of diabetes, as was the case in the Spanish National Study (7), nor was it included in predictive models of DM such as the Diabetes Risk Score(39) and the Australian Diabetes Risk Assessment (AUSDRISK)(40), although treatment of hypertension was included. However, hypertension was incorporated to the Framingham Offspring Study model(41). In addition, it was reported as an independent predictor of DM in some prospective cohort studies, such as the Women’s Health Study(42) and an 11.5-year follow-up study in the Netherlands(43). It has been postulated that an increased likelihood of insulin resistance mediates the association between hypertension and DM. The discrepancy between the different studies may be due to the degree of hypertension control of the patients included in each study, as polymorphonuclear leukocytes from patients with poor hypertension control produce significantly higher levels of O2-, hydrogen peroxide (H2O2) and lipid peroxides, indicating higher oxidative stress that may predispose to the development of DM(44,45).

Some studies have suggested that the triglyceride-glucose ratio is a surrogate marker of insulin resistance and a predictor of type 2 diabetes(46) and poor glycaemic control in overweight and obese patients(47). However, we did not find an association with the development of DM. As insulin resistance and insufficient pancreatic insulin secretion are required for the development of DM2, it is possible that the patients in our study developed DM2 primarily because of insufficient insulin secretion and not because of resistance to its peripheral action.

Furthermore, while in some studies (4,5,9) sex, age, family history of T2DM or waist circumference were identified as predictors of progression to T2DM, in our study, they were not identified as such.

### Weaknesses

In the absence of a specific date for diagnosis of T2DM, we approximated this date to be 1 June of the year in which either the blood tests or the initiation of treatment with antidiabetic drugs suggested the onset of a new case of T2DM. A single assessment of glucose status may not be sufficient to predict T2DM, given the dynamic nature of glucose metabolism. Patients with T2DM who were initially managed with diet and exercise without medication may have been missed. The possible effects of unmeasured confounding variables such as genetic predisposition or time-varying factors (changes in weight, diet, physical activity during follow-up) remain.

### Strengths

The main strength of the present study was the good characterisation of the sample, the long follow-up, the broad criteria for defining the incidence of T2DM, a rigorous selection of risk factors, including not only glucose levels and some anthropometric assessments, but also some lifestyle-related factors such as physical activity, self-perception of health and behavioural characteristics.

Compared to studies carried out in Spain, our study offers a complementary perspective with a reference population of patients from Madrid city characterised by their BGS and a survival analysis using a Cox proportional hazards explanatory model of progression to T2DM with BGS as the main variable both in the total study population and in patients with pre-T2DM (sensitivity analysis). Furthermore, since our clinical data to determine progression to T2DM were obtained from the Madrid PC-HER, no recapture of patients was necessary for the follow-up period, thereby avoiding a possible selection bias. Finally, we incorporated the patient’s perspective as an independent variable in the Cox model through the SF-36 self-perception of health questionnaire.

## Conclusion

The incidence rate of T2DM in the city of Madrid is aligned with the figures published by other studies carried out in Spain. Regarding the predictors of progression to T2DM, BGS in its isolated IGT and IFG-IGT patient categories, together with obesity, increases the risk in both NG and prediabetes patients. In turn, being on diuretic treatment in the case of persons with NG or patients with prediabetes, as well as notifying the patients with prediabetes of their condition, is a protective factor against progression to T2DM.

## Data Availability

Datasets generated and/or analysed during the current study are available from the corresponding author on reasonable request

## Acknowledgments

We would like to thank all members of the SPREDIA-2 Group: Leopoldo Pérez-Isla (Hospital Clínico de San Carlos), Ignacio Vicente (CS Monóvar), Sara Artola (CS Ma Jesús Hereza), Ma Isabel Granados-Menéndez (CS Monóvar), Domingo Beamud-Victoria (CS Felipe II), Isidoro Dujovne-Kohan (CS Los Castillos), Rosa María Chico-Moraleja (Hospital Central de la Defensa), Carmen Martín-Madrazo (CS Monóvar), Juan Cárdenas-Valladolid (Gerencia de Atención Primaria), Rosario Echegoyen de Nicolás (CS Benita de Ávila), Concepción Aguilera Linde (CS Ciudad Periodistas), Álvaro R Aguirre De Carcer Escolano (CS La Ventilla), Patricio Alonso Sacristán (CS Ciudad Periodistas), M Jesús Álvarez Otero (CS Dr Castroviejo), Paloma Arribas Pérez (CS Santa Hortensia), Maria Luisa Asensio Ruiz (CS Fuentelarreina), Pablo Astorga Díaz (CS Barrio Pilar), Begoña Berriatua Ena (CS Dr Castroviejo), Ana Isabel Bezos Varela (CS José Marva), María José Calatrava Triguero (CS Ciudad Jardín), Carlos Casanova García (CS Barrio Pilar), Ángeles Conde Llorente (CS Barrio Pilar), Concepción Díaz Laso (CS Fuentelarreina), Emilia Elviro García (CS Ciudad Periodistas), Orlando Enríquez Dueñas (CS Fuentelarreina), María Isabel Ferrer Zapata (CS El Greco), Froilán Antuña (CS Ciudad Periodistas), Maria Isabel García Lazaro (CS Ciudad Periodistas), Maria Teresa Gómez Rodríguez (CS Barrio Pilar), África Gómez Lucena (CS La Ventilla), Francisco Herrero Hernández (CS La Ventilla), Rosa Julián Viñals (CS Dr Castroviejo), Gerardo López Ruiz Ogarrio “in memoriam” (CS Barrio Pilar), Maria Del Carmen Lumbreras Manzano (CS José Marva), Sonsoles Paloma Luquero López (CS Ciudad Periodistas), Ana Martínez Cabrera Peláez (CS Barrio Pilar), Montserrat Nieto Candenas (CS La Ventilla), María Alejandra Rabanal Carrera (CS Barrio Pilar), Ángel Castellanos Rodríguez (CS Ciudad Periodistas), Ana López Castellanos (CS La Ventilla), Milagros Velázquez García (CS Barrio Pilar) and Margarita Ruiz Pacheco (CS Dr. Castroviejo)

