## Supplemental_material for "Prospective population-based observational study to estimate the incidence of T2DM in a metropolitan population in the north of Madrid (Spain) and to determine the effect of baseline glycaemic status through an explanatory Cox model. SPRINDIAP-1 study (Secondary PRevention of INcident DIAbetes in pa"

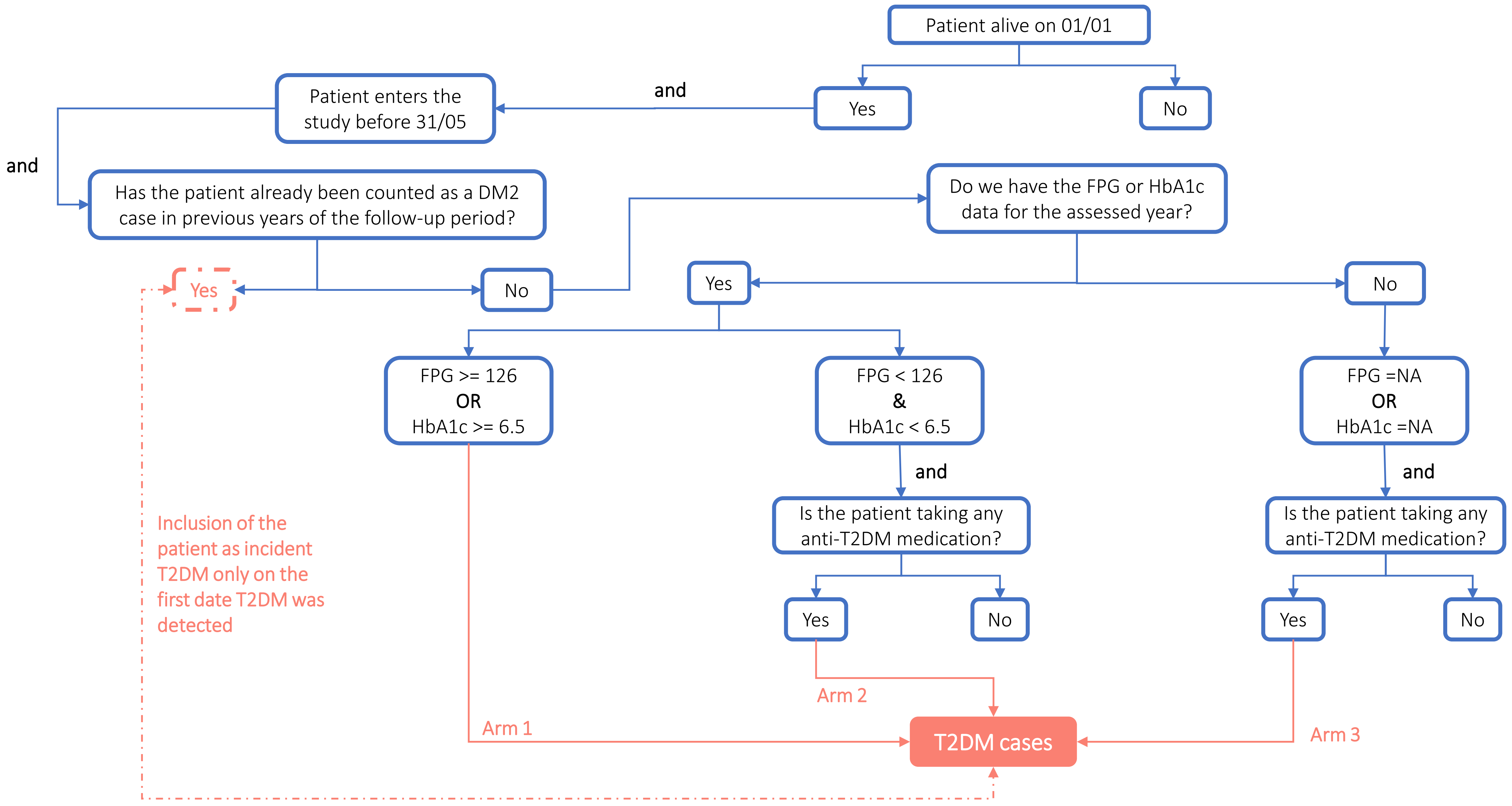


**Decision algorithm for the identification of T2DM cases year to year (2011-2019)**

| FPG | 2h-OGTT glucose | Baseline glycaemic phenotype |
|---|---|---|
| 99 | NA | Normoglycaemic |
| 93 | NA | Normoglycaemic |
| 91 | NA | Normoglycaemic |
| 92 | NA | Normoglycaemic |
| 86 | NA | Normoglycaemic |
| 96 | NA | Normoglycaemic |
| 128 | 160 | IGT |
| 144 | 174 | IGT |
| 130 | 177 | IGT |
| 133 | 141 | IGT |
| 145 | 156 | IGT |
| 130 | 191 | IGT |
| 133 | 177 | IGT |
| 127 | 154 | IGT |
| 127 | 168 | IGT |
| 129 | 157 | IGT |
| 127 | 167 | IGT |
| 131 | 153 | IGT |
| 126 | 160 | IGT |

| FPG | 2h-OGTT glucose | Baseline glycaemic phenotype |
|---|---|---|
| 126 | 72 | IFG |
| 138 | 119 | IFG |
| 100 | NA | IFG |
| 133 | 135 | IFG |
| 127 | 67 | IFG |
| 107 | NA | IFG |
| 128 | 133 | IFG |
| 126 | 116 | IFG |
| 103 | NA | IFG |
| 110 | NA | IFG |
| 118 | NA | IFG |
| 126 | 135 | IFG |
| 126 | 125 | IFG |
| 103 | NA | IFG |
| 110 | NA | IFG |
| 132 | 69 | IFG |
| 126 | 113 | IFG |

**Patients whose baseline glycaemic phenotype was decided on a case-by-case basis by the study Steering Committee as they could not be categorised according to the American Diabetes Association (ADA) criteria**. FPG: Fasting plasma glucose; 2h-OGTT glucose: venous blood plasma glucose at two hours after the 75 g Oral Glucose Overload

### Sex and age distribution of the baseline study population stratified by baseline glycaemic status

| Basal glycemic status | | Sex | Under 55 years old | | 55-64 years old | | 65 and over | |
|---|---|---|---|---|---|---|---|---|
| | | | n | Percentage (95% CI) | n | Percentage (95% CI) | n | Percentage (95% CI) |
| Non-preDM2 | NGT patients | Male | 40 | 3.28 (2.35-4.41) | 69 | 5.66 (4.43-7.11) | 40 | 3.28 (2.35-4.41) |
| | | Female | 77 | 6.31 (5.02–7.83) | 209 | 17.15 (15.07-19.38) | 125 | 10.25 (8.61-12.10) |
| PreDM2 | IFG patients | Male | 39 | 3.19 (2.28-4.35) | 105 | 8.61 (7.10-10.33) | 72 | 5.91 (4.65-7.38) |
| | | Female | 20 | 1.64 (1.00-2.52) | 114 | 9.35 (7.78-11.13) | 68 | 5.58 (4.36–7.02) |
| | IGT patients | Male | 4 | 0.33 (0.09-0.84) | 17 | 1.39 (0.81-2.22) | 14 | 1.14 (0.63-1.92) |
| | | Female | 1 | 0.08 (0.00-0.46) | 15 | 1.23 (0.69-2.02) | 19 | 1.56 (0.94-2.42) |
| | IFG-IGT patients | Male | º | 0.82 (0.39-1.50) | 36 | 2.95 (2.07-4.06) | 39 | 1.15 (0.62-1.92) |
| | | Female | 7 | 0.08 (0.00-0.45) | 38 | 3.11 (2.22-4.25) | 41 | 3.36 (2.42-4.53) |

IFG: Isolated impaired fasting glucose patients; IGT: Isolated impaired glucose tolerance patients;
IFG-IGT: patients with IFG and IGT; NGT: Normal glucose tolerance patients; pre-DM2: prediabetic patients

### Raw T2DM rates by baseline glycaemic phenotype

| Baseline glycaemic phenotype categories (n) | T2DM | | | | |
|---|---|---|---|---|---|
| | Cases | Persons-year | Inc rate/1.000 persons-years (CI 95%) | Inc rate ratio (CI 95%*) | p-value** of rate comparison |
| All (1,219) | 97 | 8651 | 11.21 (9.09 - 13.68) | | |
| NGT (560) | 23 | 4105 | 5.60 (3,55 - 8,41) | 1 | - |
| Pre-DM2 (659) | 74 | 4546 | 16.28 (12.78% - 20.43%) | 2.91 (1.80 - 4.86) | < 0.01 |
| IFG (418) | 32 | 2989 | 10.71 (7,32 - 15,11) | 1.91 (1.08 - 3.42) | < 0.05 |
| IGT (70) | 12 | 446 | 26.91 (13,90 - 47,00) | 4.80 (2.18 -10.06) | < 0.01 |
| IFG-IGT (171) | 30 | 1111 | 27.00 (18,22 - 38,55) | 4.82 (2.71 - 8.69) | < 0.01 |

* as calculated by Rothman's exact method; ** as calculated by Rosner's rate-comparison test

IFG: Isolated impaired fasting glucose patients; IGT: Isolated impaired glucose tolerance patients; IFG-IGT: patients with IFG and IGT; IR: incidence rate; NGT: Normal glucose tolerance patients; pre-DM2: Pre-diabetic patients; T2DM: Type 2 Diabetes Mellitus

### Raw T2DM rates by sex

| Sex (n) | Cases | Persons-year | Inc rate/1.000 persons-years (CI 95%) | Inc rate ratio (CI 95%*) | p-value** of rate comparison |
|---|---|---|---|---|---|
| Women (734) | 52 | 5295 | 9.82 (7.33 - 12.88) | 1 | - |
| Men (485) | 45 | 3357 | 13.40 (9.78 - 17.94) | 1.36 (0.90 - 2.07) | > 0.05 |

* as calculated by Rothman's exact method; ** as calculated by Rosner's rate-comparison test

### Raw T2DM rates by age

| Age (n) | Cases | Persons-year | Inc rate/1.000 persons-years (CI 95%) | Inc rate ratio (CI 95%*) | p-value** of rate comparison |
|---|---|---|---|---|---|
| < 55 years old (198) | 11 | 1446 | 7.61 (3.80 - 13.61) | 1 | - |
| 55-64 years old (603) | 50 | 4276 | 11.69 (8.68 - 15.42) | 1.54 (0.79 - 3.27) | > 0.05 |
| > 65 years old (418) | 36 | 2929 | 12.29 (8.61 - 17.01) | 1.62 (0.81 - 3.52) | > 0.05 |

* as calculated by Rothman's exact method; ** as calculated by Rosner's rate-comparison test

### Raw T2DM rates by baseline glycaemic status and age group in males and females

| Impaired fasting glucose (IFG) vs Normal glucose tolerance (NGT) | | IFG at baseline (n= 418) | | | | NGT at baseline (n=560) | | | | | |
|---|---|---|---|---|---|---|---|---|---|---|---|
| | | n | T2DM cases | p-y | T2DM incidence per 1000 p-y (95% CI) | n | T2DM cases | p-y | T2DM incidence per 1000 p-y (95% CI) | Raw rate ratio (IFG/NGT) (95% CI) | p-value |
| **All** | < 55 years old | 59 | 2 | 430.80 | 4.64 (0.56-16.77) | 117 | 5 | 878.40 | 5.69 (1.85-13.28) | 0.81 (0.08-4.98) | 0.852 |
| | 55-64 years old | 219 | 17 | 1581.00 | 10.75 (6.26-17.22) | 278 | 9 | 2025.70 | 4.44 (2.03-8.43) | 2.42 (1.02 -6.16) | 0.030 |
| | > 65 years old | 140 | 13 | 977.50 | 13.30 (7.08-22.74) | 165 | 9 | 1201.00 | 7.49 (3.43-14.23) | 1.78 (0.70-4.71) | 0.190 |
| | **All** | **418** | **32** | **2989.3** | **10.73 (7.34-15.14)** | **560** | **23** | **4105.1** | **5.60 (3.55-8.41)** | **1.91 (1.08-3.42)** | **0.018** |
| **Men** | < 55 years old | 39 | 2 | 275.80 | 7.25 (0.88-26.20) | 40 | 1 | 306.60 | 3.26 (0.08-18.17) | 2.22 (0.12-131.17) | 0.567 |
| | 55-64 years old | 105 | 9 | 740.70 | 12.15 (5.56-23.07) | 69 | 1 | 495.70 | 2.02 (0.05-11.24) | 0.02 (0.83-264.00) | 0,052 |
| | > 65 years old | 72 | 4 | 516.50 | 7.74 (2.11-19.83) | 40 | 4 | 269.40 | 14.85 (4.05-38.02) | 0.52 (0.10-2.80) | 0.376 |
| | **All** | **216** | **15** | **1533** | **9.79 (5.48-16.14)** | **149** | **6** | **1071.7** | **5.60 (2.06-12.19)** | **1.75 (0.64-5.50)** | **0.251** |
| **Women** | < 55 years old | 20 | 0 | 155.00 | 0.00 | 77 | 4 | 571.80 | 6.96 (1.91-17.91) | - | - |
| | 55-64 years old | 114 | 8 | 840.30 | 9.52 (4.11-18.76) | 209 | 8 | 1530.00 | 5.23 (2.26-20.30) | 1.82 (0.60-5.57) | 0.241 |
| | > 65 years old | 68 | 9 | 461.00 | 19.52 (8.93-37.06) | 125 | 5 | 931.60 | 5.37 (1.74-12.53) | 3.64 (1.10-13.82) | 0.020 |
| | **All** | **202** | **17** | **1456.3** | **11.68 (6.80-18.69)** | **411** | **17** | **3033.4** | **5.60 (3.27-8.97)** | **2.08 (1.00-4.34)** | **0.036** |

IFG: Isolated impaired fasting glucose patients; NGT: Normal glucose tolerance patients

| Impaired glucose tolerance (IGT) vs Normal glucose tolerance (NGT) | | | | | | | | | | | |
|---|---|---|---|---|---|---|---|---|---|---|---|
| | | IGT at baseline (n=70 ) | | | | NGT at baseline (n=560) | | | | | |
| | | n | T2DM cases | p-y | T2DM incidence per 1000 p-y (95% CI) | n | T2DM cases | p-y | T2DM incidence per 1000 p-y (95% CI) | Raw rate ratio (IGT/NGT) (95% CI) | p-value |
| All | < 55 years old | 5 | 2 | 23.63 | 84.64 (10.25-305.74) | 117 | 5 | 878.40 | 5.69 (1.85-13.28) | 14.87 (1.45-90.82) | 0.014 |
| | 55-64 years old | 32 | 4 | 215.90 | 18.53 (50.05-47.44) | 278 | 9 | 2025.70 | 4.44 (20.3-8.43) | 4.17 (0.94-14.94) | 0.036 |
| | > 65 years old | 33 | 6 | 206.50 | 29.06 (10.66-63.24) | 165 | 9 | 1201.00 | 7.49 (3.43-14.23) | 3.88 (1.14-12.20) | 0.018 |
| | **All** | **70** | **12** | **446.03** | **26.90 (13.90-47.00)** | **560** | **23** | **4105.1** | **5.60 (3.55-8.41)** | **4.80 (2.18-10.56)** | **0.000** |
| Men | < 55 years old | 4 | 2 | 17.42 | 114.81 (13.90-414-74) | 40 | 1 | 306.60 | 3.26 (0.08-18.17) | 35.20 (1.83-2076.76) | 0.009 |
| | 55-64 years old | 17 | 3 | 111.90 | 26.81 (5.53-78.35) | 69 | 1 | 495.70 | 2.02 (0.05-11.24) | 13.29 (1.07-697.66) | 0.023 |
| | > 65 years old | 14 | 5 | 73.90 | 67.66 (21.97-157-89) | 40 | 4 | 269.40 | 14.85 (4.05-38.02) | 4.56 (0.98-22.97) | 0.031 |
| | **All** | **35** | **10** | **203.22** | **49.20 (23.60-90.50)** | **149** | **6** | **1071.7** | **5.60 (2.06-12.09)** | **8.79 (2.90-29.43)** | **0.000** |
| Women | < 55 years old | 1 | 0 | 6.21 | 0.00 | 77 | 4 | 571.80 | 7.00 (1.91-17.91) | - | - |
| | 55-64 years old | 15 | 1 | 104.00 | 9.62 (0.24-53.57) | 209 | 8 | 1530.00 | 5.23 (2.26-10.30) | 1.84 (0.04-13.72) | 0.555 |
| | > 65 years old | 19 | 1 | 132.60 | 7.54 (0.19-42.02) | 125 | 5 | 931.60 | 5.37 (1.74-12.53) | 1.41 (0.03-12.56) | 0.716 |
| | **All** | **35** | **2** | **242.81** | **8.23 (1.00-29.75)** | **411** | **17** | **3033.4** | **5.6 (3.27-8.97)** | **1.47 (0.17-6.19)** | **0.579** |

IGT: Isolated impaired glucose tolerance patients; NGT: Normal glucose tolerance patients

| Impaired fasting glucose & Impaired glucose tolerance (IFG-IGT) vs Normal glucose tolerance (NGT) | | | | | | | | | | | |
|---|---|---|---|---|---|---|---|---|---|---|---|
| | | IFG-IGT at baseline (n= 171) | | | | NGT at baseline (n=560) | | | | | |
| | | n | T2DM cases | p-y | T2DM incidence per 1000 p-y (95% CI) | n | T2DM cases | p-y | T2DM incidence per 1000 p-y (95% CI) | Raw rate ratio (IFG-IGT/NGT) (95% CI) | p-value |
| **All** | < 55 years old | 17 | 2 | 113.19 | 17.67 (2.14-63.89) | 117 | 5 | 878.40 | 5.69 (1.85-13.28) | 3.10 (0.30-18.96) | 0.222 |
| | 55-64 years old | 74 | 20 | 453.60 | 44.09 (26.93-68.96) | 278 | 9 | 2025.70 | 4.44 (2.03-8.43) | 9.92 (4.32-24.75) | 0.000 |
| | > 65 years old | 80 | 8 | 544.30 | 14.70 (6.35-28.96) | 165 | 9 | 1201.00 | 7.49 (3.43-14.23) | 1.96 (0.66-5.73) | 0.177 |
| | **All** | **171** | **30** | **1111.09** | **27.01 (18.22-38.55)** | **560** | **23** | **4105.1** | **5.60 (3.55-8.41)** | **4.82 (2.71-8.67)** | **0.000** |
| **Men** | < 55 years old | 10 | 1 | 67.66 | 14.78 (0.37-82.35) | 40 | 1 | 306.60 | 3.26 (0.08-18.17) | 4.5 (0.06-355.71) | 0.367 |
| | 55-64 years old | 36 | 9 | 229.10 | 39.28 (17.96-74.57) | 69 | 1 | 495.70 | 2.02 (0.05-11.24) | 19.47 (2.70-853-53) | 0.000 |
| | > 65 years old | 39 | 4 | 252.00 | 15.87 (4.33-40.64) | 40 | 4 | 269.40 | 14.85 (4.05-38.02) | 1.07 (0.20-5.74) | 0.927 |
| | **All** | **85** | **14** | **548.76** | **25.51 (13.95-42.80)** | **149** | **6** | **1071.7** | **5.60 (2.06-12.19)** | **4.56 (1.65-14.46)** | **0.001** |
| **Women** | < 55 years old | 7 | 1 | 45.53 | 21.96 (0.56-122.37) | 77 | 4 | 571.80 | 7.00 (1.91-27.91) | 3.14 (0.06-31.73) | 0.365 |
| | 55-64 years old | 38 | 11 | 224.50 | 49.00 (24.46-87.67) | 209 | 8 | 1530.00 | 5.23 (2.26-10.30) | 9.37 (3.43-26.84) | 0.000 |
| | > 65 years old | 41 | 4 | 292.30 | 13.69 (3.73-35.04) | 125 | 5 | 931.60 | 5.37 (1.74-12.53) | 2.55 (0.51-11.85) | 0.186 |
| | **All** | **86** | **16** | **562.33** | **28.45 (16.26-46.21)** | **411** | **17** | **3033.4** | **5.60 (2.37-8.97)** | **5.07 (2.40-10.69)** | **0.000** |

IFG-IGT: patients with IFG and IGT; NGT: Normal glucose tolerance patients

### Raw and Standardized T2DM rates by baseline glycaemic phenotype and age group in males and females (PRE-DM2 vs NGT)

| | | pre-DM2 | | | | NGT | | | | Raw rate ratio (pre-DM2/NG) | Standardized rate | | | |
|---|---|---|---|---|---|---|---|---|---|---|---|---|---|---|
| | | n | T2DM cases | p-y | T2DM raw IR /1000 p-y | n | T2DM cases | p-y | T2DM raw IR /1000 p-y | | Weight (p-y) | T2DM IR /1000 p-y (pre-DM2) | T2DM IR /1000 p-y (NG) | Rate ratio (Pre-DM2/NG) |
| **All** | Under 55 years old | 81 | 6 | 567.62 | 10.57 | 117 | 5 | 878.40 | 5.69 | 1.86 | 1446.02 | | | |
| | 55 to 64 years old | 325 | 41 | 2250.50 | 18.22 | 278 | 9 | 2025.70 | 4.44 | 4.10 | 4276.2 | 16.1 | 5.7 | **2.83** |
| | 65 years old or older | 253 | 27 | 1728.30 | 15.62 | 165 | 9 | 1201.00 | 7.49 | 2.08 | 2929.3 | | | |
| | | **659** | **74** | **4546.4** | **16.3** | **560** | **23** | **4105.1** | **5.6** | **2.91** | **8651.52** | | | |

| | | pre-DM2 | | | | NGT | | | | Raw rate ratio (pre-DM2/NG) | Standardized rate | | | |
|---|---|---|---|---|---|---|---|---|---|---|---|---|---|---|
| | | n | T2DM cases | p-y | T2DM raw IR /1000 p-y | n | T2DM cases | p-y | T2DM raw IR /1000 p-y | | Weight (p-y) | T2DM IR /1000 p-y (pre-DM2) | T2DM IR /1000 p-y (NG) | Rate ratio (Pre-DM2/NG) |
| **Men** | Under 55 years old | 53 | 5 | 360.88 | 13.86 | 40 | 1 | 306.60 | 3.26 | 4.25 | 667.5 | | | |
| | 55 to 64 years old | 158 | 21 | 1081.70 | 19.41 | 69 | 1 | 495.70 | 2.02 | 9.62 | 1577.4 | 17.0 | 6.5 | **2.61** |
| | 65 years old or older | 125 | 13 | 842.40 | 15.43 | 40 | 4 | 269.40 | 14.85 | 1.04 | 1111.8 | | | |
| | | **336** | **39** | **2285** | **17.1** | **149** | **6** | **1071.7** | **5.6** | **3.05** | **3356.68** | | | |

| | | pre-DM2 | | | | NGT | | | | Raw rate ratio (pre-DM2/NG) | Standardized rate | | | |
|---|---|---|---|---|---|---|---|---|---|---|---|---|---|---|
| | | n | T2DM cases | p-y | T2DM raw IR /1000 p-y | n | T2DM cases | p-y | T2DM raw IR /1000 p-y | | Weight (p-y) | T2DM IR /1000 p-y (pre-DM2) | T2DM IR /1000 p-y (NG) | Rate ratio (Pre-DM2/NG) |
| **Women** | Under 55 years old | 28 | 1 | 206.74 | 4.84 | 77 | 4 | 571.80 | 7.00 | 0.69 | 778.54 | | | |
| | 55 to 64 years old | 167 | 20 | 1168.80 | 17.11 | 209 | 8 | 1530.00 | 5.23 | 3.27 | 2698.8 | 14.9 | 5.5 | **2.68** |
| | 65 years old or older | 128 | 14 | 885.90 | 15.80 | 125 | 5 | 931.60 | 5.37 | 2.94 | 1817.5 | | | |
| | | **323** | **35** | **2261.4** | **15.5** | **411** | **17** | **3033.4** | **5.6** | **2.76** | **5294.84** | | | |

IR: incidence rate; NGT:Normal glucose tolerance patients; pre-DM2: Pre-diabetic patients; p-y: persons-year; T2DM: Type 2 Diabetes Mellitus

T2DM raw IR /1000 p-y: Ratio between T2DM cases and p-y; Weight: Sum of persons year at risk (pre-DM2 + NGT)

T2DM IR /1000 p-y: Weighted average of raw T2DM raw IR /1000 p-y rates in each baseline glycaemic phenotype stratum

T2DM IR /1000 p-y: Weighted average of raw T2DM raw IR /1000 p-y rates in each baseline glycaemic phenotype stratum

### IFG vs NGT

| All | | IFG | | | | NGT | | | | Raw rate ratio (IFG/NG) | Standardized rate | | | |
|---|---|---|---|---|---|---|---|---|---|---|---|---|---|---|
| | | n | T2DM cases | p-y | T2DM raw IR /1000 p-y | n | T2DM cases | p-y | T2DM raw IR /1000 p-y | | Weight (p-y) | T2DM IR /1000 p-y (IFG) | T2DM IR /1000 p-y (NG) | Rate ratio (Pre-DM2/NG) |
| **All** | Under 55 years old | 59 | 2 | 430.80 | 4.64 | 117 | 5 | 878.40 | 5.69 | 0.82 | 1309.2 | 10.4 | 5.6 | **1.86** |
| | 55 to 64 years old | 219 | 17 | 1581.00 | 10.75 | 278 | 9 | 2025.70 | 4.44 | 2.42 | 3606.7 | | | |
| | 65 years old or older | 140 | 13 | 977.50 | 13.30 | 165 | 9 | 1201.00 | 7.49 | 1.77 | 2178.5 | | | |
| | | **418** | **32** | **2989.3** | **10.7** | **560** | **23** | **4105.1** | **5.6** | **1.91** | **7094.40** | | | |

| | | IFG | | | | NGT | | | | Raw rate ratio (IFG/NG) | Standardized rate | | | |
|---|---|---|---|---|---|---|---|---|---|---|---|---|---|---|
| | | n | T2DM cases | p-y | T2DM raw IR /1000 p-y | n | T2DM cases | p-y | T2DM raw IR /1000 p-y | | Weight (p-y) | T2DM IR /1000 p-y (IFG) | T2DM IR /1000 p-y (NG) | Rate ratio (Pre-DM2/NG) |
| **Men** | Under 55 years old | 39 | 2 | 275.80 | 7.25 | 40 | 1 | 306.60 | 3.26 | 2.22 | 582.4 | 9.7 | 6.2 | **1.58** |
| | 55 to 64 years old | 105 | 9 | 740.70 | 12.15 | 69 | 1 | 495.70 | 2.02 | 6.02 | 1236.4 | | | |
| | 65 years old or older | 72 | 4 | 516.50 | 7.74 | 40 | 4 | 269.40 | 14.85 | 0.52 | 785.9 | | | |
| | | **216** | **15** | **1533** | **9.8** | **149** | **6** | **1071.7** | **5.6** | **1.75** | **2604.70** | | | |

| | | IFG | | | | NGT | | | | Raw rate ratio (IFG/NG) | Standardized rate | | | |
|---|---|---|---|---|---|---|---|---|---|---|---|---|---|---|
| | | n | T2DM cases | p-y | T2DM raw IR /1000 p-y | n | T2DM cases | p-y | T2DM raw IR /1000 p-y | | Weight (p-y) | T2DM IR /1000 p-y (IFG) | T2DM IR /1000 p-y (NG) | Rate ratio (Pre-DM2/NG) |
| **Women** | Under 55 years old | 20 | 0 | 155.00 | 0.00 | 77 | 4 | 571.80 | 7.00 | 0.00 | 726.8 | 11.1 | 5.6 | **1.99** |
| | 55 to 64 years old | 114 | 8 | 840.30 | 9.52 | 209 | 8 | 1530.00 | 5.23 | 1.82 | 2370.3 | | | |
| | 65 years old or older | 68 | 9 | 461.00 | 19.52 | 125 | 5 | 931.60 | 5.37 | 3.64 | 1392.6 | | | |
| | | **202** | **17** | **1456.3** | **11.7** | **411** | **17** | **3033.4** | **5.6** | **2.08** | **4489.70** | | | |

IFG: Isolated impaired fasting glucose patients; IR: incidence rate; NGT:Normal glucose tolerance patients; pre-DM2: Pre-diabetic patients; p-y: persons-year;
T2DM: Type 2 Diabetes Mellitus; T2DM raw IR /1000 p-y: Ratio between T2DM cases and p-y; Weight: Sum of persons year at risk (IFG + NGT)
T2DM IR /1000 p-y: Weighted average of raw T2DM raw IR /1000 p-y rates in each baseline glycaemic phenotype stratum

**IGT vs NGT**

| | | IGT | | | | NGT | | | | Raw rate ratio (IGT/NG) | Standardized rate | | | |
|---|---|---|---|---|---|---|---|---|---|---|---|---|---|---|
| | | n | T2DM cases | p-y | T2DM raw IR /1000 p-y | n | T2DM cases | p-y | T2DM raw IR /1000 p-y | | Weight (p-y) | T2DM IR /1000 p-y (IGT) | T2DM IR /1000 p-y (NG) | Rate ratio (Pre-DM2/NG) |
| **All** | Under 55 years old | 5 | 2 | 23.63 | 84.64 | 117 | 5 | 878.40 | 5.69 | 14.87 | 902.03 | 34.9 | 5.6 | **6.19** |
| | 55 to 64 years old | 32 | 4 | 215.90 | 18.53 | 278 | 9 | 2025.70 | 4.44 | 4.17 | 2241.6 | | | |
| | 65 years old or older | 33 | 6 | 206.50 | 29.06 | 165 | 9 | 1201.00 | 7.49 | 3.88 | 1407.5 | | | |
| | | **70** | **12** | **446.03** | **26.9** | **560** | **23** | **4105.1** | **5.6** | **4.80** | **4551.13** | | | |

| | | IGT | | | | NGT | | | | Raw rate ratio (IGT/NG) | Standardized rate | | | |
|---|---|---|---|---|---|---|---|---|---|---|---|---|---|---|
| | | n | T2DM cases | p-y | T2DM raw IR /1000 p-y | n | T2DM cases | p-y | T2DM raw IR /1000 p-y | | Weight (p-y) | T2DM IR /1000 p-y (IGT) | T2DM IR /1000 p-y (NG) | Rate ratio (Pre-DM2/NG) |
| **Men** | Under 55 years old | 4 | 2 | 17.42 | 114.81 | 40 | 1 | 306.60 | 3.26 | 35.20 | 324.02 | 60.2 | 5.8 | **10.40** |
| | 55 to 64 years old | 17 | 3 | 111.90 | 26.81 | 69 | 1 | 495.70 | 2.02 | 13.29 | 607.6 | | | |
| | 65 years old or older | 14 | 5 | 73.90 | 67.66 | 40 | 4 | 269.40 | 14.85 | 4.56 | 343.3 | | | |
| | | **35** | **10** | **203.22** | **49.2** | **149** | **6** | **1071.7** | **5.6** | **8.79** | **1274.92** | | | |

| | | IGT | | | | NGT | | | | Raw rate ratio (IGT/NG) | Standardized rate | | | |
|---|---|---|---|---|---|---|---|---|---|---|---|---|---|---|
| | | n | T2DM cases | p-y | T2DM raw IR /1000 p-y | n | T2DM cases | p-y | T2DM raw IR /1000 p-y | | Weight (p-y) | T2DM IR /1000 p-y (IGT) | T2DM IR /1000 p-y (NG) | Rate ratio (Pre-DM2/NG) |
| **Women** | Under 55 years old | 1 | 0 | 6.21 | 0.00 | 77 | 4 | 571.80 | 7.00 | 0.00 | 578.01 | 7.2 | 5.6 | **1.30** |
| | 55 to 64 years old | 15 | 1 | 104.00 | 9.62 | 209 | 8 | 1530.00 | 5.23 | 1.84 | 1634.0 | | | |
| | 65 years old or older | 19 | 1 | 132.60 | 7.54 | 125 | 5 | 931.60 | 5.37 | 1.41 | 1064.2 | | | |
| | | **35** | **2** | **242.81** | **8.2** | **411** | **17** | **3033.4** | **5.6** | **1.47** | **3276.21** | | | |

IGT: Isolated impaired glucose tolerance patients; IR: incidence rate; NGT:Normal glucose tolerance patients; pre-DM2: Pre-diabetic patients; p-y: persons-year;
T2DM: Type 2 Diabetes Mellitus;T2DM raw IR /1000 p-y: Ratio between T2DM cases and p-y; Weight: Sum of persons year at risk (IGT + NG)
T2DM IR /1000 p-y: Weighted average of raw T2DM raw IR /1000 p-y rates in each baseline glycaemic phenotype stratum

| IFG-IGT vs NGT | | | | | | | | | | | | | | |
|---|---|---|---|---|---|---|---|---|---|---|---|---|---|---|
| | | IFG-IGT | | | | NGT | | | | | Standardized rate | | | |
| | | n | T2DM cases | p-y | T2DM raw IR /1000 p-y | n | T2DM cases | p-y | T2DM raw IR /1000 p-y | Raw rate ratio (IFG-IGT/NG) | Weight (p-y) | T2DM IR /1000 p-y (IFG-IGT) | T2DM IR /1000 p-y (NG) | Rate ratio (Pre-DM2/NG) |
| **All** | Under 55 years old | 17 | 2 | 113.19 | 17.67 | 117 | 5 | 878.40 | 5.69 | 3.10 | 991.59 | 29.2 | 5.7 | **5.13** |
| | 55 to 64 years old | 74 | 20 | 453.60 | 44.09 | 278 | 9 | 2025.70 | 4.44 | 9.92 | 2479.3 | | | |
| | 65 years old or older | 80 | 8 | 544.30 | 14.70 | 165 | 9 | 1201.00 | 7.49 | 1.96 | 1745.3 | | | |
| | | **171** | **30** | **1111.1** | **27.0** | **560** | **23** | **4105.1** | **5.6** | **4.82** | **5216.19** | | | |

| | | IFG-IGT | | | | NGT | | | | | Standardized rate | | | |
|---|---|---|---|---|---|---|---|---|---|---|---|---|---|---|
| | | n | T2DM cases | p-y | T2DM raw IR /1000 p-y | n | T2DM cases | p-y | T2DM raw IR /1000 p-y | Raw rate ratio (IFG-IGT/NG) | Weight (p-y) | T2DM IR /1000 p-y (IFG-IGT) | T2DM IR /1000 p-y (NG) | Rate ratio (Pre-DM2/NG) |
| **Men** | Under 55 years old | 10 | 1 | 67.66 | 0.00 | 40 | 1 | 306.60 | 3.26 | 0.00 | 374.26 | 22.7 | 6.4 | **3.53** |
| | 55 to 64 years old | 36 | 9 | 229.10 | 39.28 | 69 | 1 | 495.70 | 2.02 | 19.47 | 724.8 | | | |
| | 65 years old or older | 39 | 4 | 252.00 | 15.87 | 40 | 4 | 269.40 | 14.85 | 1.07 | 521.4 | | | |
| | | **85** | **14** | **548.76** | **25.5** | **149** | **6** | **1071.7** | **5.6** | **4.56** | **1620.46** | | | |

| | | IFG-IGT | | | | NGT | | | | | Standardized rate | | | |
|---|---|---|---|---|---|---|---|---|---|---|---|---|---|---|
| | | n | T2DM cases | p-y | T2DM raw IR /1000 p-y | n | T2DM cases | p-y | T2DM raw IR /1000 p-y | Raw rate ratio (IFG-IGT/NG) | Weight (p-y) | T2DM IR /1000 p-y (IFG-IGT) | T2DM IR /1000 p-y (NG) | Rate ratio (Pre-DM2/NG) |
| **Women** | Under 55 years old | 7 | 1 | 45.53 | 21.96 | 77 | 4 | 571.80 | 7.00 | 3.14 | 617.33 | 32.3 | 5.6 | **5.80** |
| | 55 to 64 years old | 38 | 11 | 224.50 | 49.00 | 209 | 8 | 1530.00 | 5.23 | 9.37 | 1754.5 | | | |
| | 65 years old or older | 41 | 4 | 292.30 | 13.68 | 125 | 5 | 931.60 | 5.37 | 2.55 | 1223.9 | | | |
| | | **86** | **16** | **562.33** | **28.5** | **411** | **17** | **3033.4** | **5.6** | **5.08** | **3595.73** | | | |

IFG-IGT: patients with IFG and IGT; IR: incidence rate; NGT:Normal glucose tolerance patients; pre-DM2: Pre-diabetic patients; p-y: persons-year; T2DM: Type 2 Diabetes Mellitus; T2DM raw IR /1000 p-y: Ratio between T2DM cases and p-y; Weight: Sum of persons year at risk (IFG-IGT + NG) T2DM IR /1000 p-y: Weighted average of raw T2DM raw IR /1000 p-y rates in each baseline glycaemic phenotype stratum
